# Hospital burden of chronic obstructive pulmonary disease exacerbation in Hong Kong in the post-COVID pandemic era – Impact of phenotype and exacerbation history

**DOI:** 10.1101/2025.04.11.25325630

**Authors:** Wang Chun Kwok, Ali Choo, Lu Zhou, Ting Fung Ma, Isaac Sze Him Leung, Chun Ka Wong, James Chung Man Ho

## Abstract

**Introduction:** Chronic obstructive pulmonary disease (COPD) has been reported to have significant healthcare and economic burdens worldwide. Since the COVID-19 pandemic, there have been changes in the epidemiology of various chronic diseases. The healthcare and economic burden of severe acute exacerbation COPD (AECOPD) and the factors affecting the burden should be assessed in the post-pandemic era.

**Methods:** A territory-wide study was conducted in Hong Kong to study the healthcare and economic burden of severe AECOPD from 2022 to 2024, and identify factors associated with an increase in the direct healthcare costs related to severe AECOPD. Adult patients with severe AECOPD admitted to the acute medical unit managed by the Hong Kong Hospital Authority from 2022 to 2024 were included. The primary outcome was the total headcount and admission numbers related to severe AECOPD, as well as the direct healthcare costs associated with severe AECOPD. The secondary outcomes included factors associated with increased direct healthcare costs resulting from severe AECOPD.

**Results:** From 2022 to 2024, there were 11,465 COPD patients admitted to acute medical hospitals for severe AECOPD, contributing to 25,053 hospital admissions and 150,717 bed days. The total direct healthcare cost was HKD $1.259 billion for these severe AECOPD. There was a significant increase in the severe AECOPD admission numbers over the 3 years. While the phenotype by baseline BEC did not affect the annual total healthcare costs related to severe AECOPD, patients with co-existing bronchiectasis had significantly higher annual number of severe AECOPD and annual direct healthcare costs. The annual number of severe AECOPD significantly correlates with total direct healthcare costs and annual direct healthcare costs related to severe AECOPD.

**Conclusions:** Severe AECOPD carries a significant burden towards healthcare system. Patients with bronchiectasis and more frequent severe AECOPD were factors associated with a higher number of severe AECOPD episodes and the associated direct healthcare costs.

## Introduction

Chronic obstructive pulmonary disease (COPD) is a common chronic respiratory disease worldwide, affecting 174.5 million people as of 2015 (1). COPD is also a prevalent disease in Hong Kong, resulting in a significant healthcare burden. It was first reported that the crude mortality rate and crude hospitalization rate were 29.1/100,000 and193/100,000, respectively, in 2005(2). Another local study revealed that in the year 2014, there were 9,613 and 19,771 total patient headcounts and admission numbers for COPD (3).

COPD contributed to a significant healthcare and economic burden, especially among moderate-to-very severe COPD(4). In European countries, the estimated per patient per year direct cost in Norway, Denmark, Germany, Italy, Sweden, Greece, Belgium, and Serbia was €10,701, 9,580, 7,847, 7,448, 7,045, 2,896, 1,963, and 2,047 (HKD$ 98,319.20, 88,256, 72,148.40, 68,569.60, 64,894, 26,675.20, 18,055.60 and 18,858.40) respectively (5). A modeling study reported that COPD will cost the world economy INT$4.326 trillion (HKD$45.45 trillion) (uncertainty interval 3·327-5·516; at constant 2017 prices) in 2020-50. This economic effect is equivalent to a yearly tax of 0·111% (0·085-0·141) on global gross domestic product (GDP). China and the USA face the largest economic burdens from COPD, accounting for INT$1·363 trillion (HKD$14.35 trillion) (uncertainty interval 1·034-1·801) and INT$1·037 trillion (HKD$10.92 trillion) (0·868-1·175), respectively(6). It is also projected that in the USA, the 20-year discounted direct medical costs attributable to COPD from 2019 to 20138 were estimated to be $800.90 billion (7). While it is known that COPD contributed to substantial healthcare costs, it is also important to know which factors contributed more to the costs. Factors reported to be associated with increasing cost of COPD management include late diagnosis, severity of disease, frequency of exacerbation, hospital readmissions, non-adherence to the therapy and exposure to COPD risk factors (5). Medicare patients with recent moderate or severe exacerbations, or at least two exacerbations per year were at significant risk for future exacerbations and incur higher all-cause costs (8).

Apart from the economic burden by healthcare utilization(9), acute exacerbation of COPD(AECOPD) is also associated with excess patient morbidity and mortality (10, 11, 12). However, between 2020 and 2021, the coronavirus disease 2019 (COVID-19) pandemic led to major changes in disease epidemiology, including in COPD, with a reduction in the hospitalization numbers (13, 14, 15).

It is important to revisit the economic burden related to COPD, especially the direct healthcare costs associated with severe AECOPD in the post-COVID era, and also evaluate the disease factors affecting healthcare costs, which may hint the clinicians on how appropriate personalized therapy may benefit the patients in both clinical and health economic aspects.

## Methods

This is a territory-wide study conducted in Hong Kong to study the burden of severe AECOPD from the year 2022 to 2024, and identify factors associated with an increase in the direct healthcare costs associated with severe AECOPD. Adult patients with severe AECOPD admitted to the acute medical unit managed by the Hong Kong Hospital Authority (HKHA) from 2022 to 2024 were included. This study utilized electronic health records from the Clinical Data Analysis and Reporting System (CDARS) managed by the HKHA. HKHA is a public healthcare service provider that operates 43 hospitals and institutions, and 122 outpatient clinics, covering more than 90% of the Hong Kong population since 1993 (16). The CDARS captures medical information, including diagnosis, drug prescription details, demographics, admissions, medical procedures, and laboratory results. The COPD diagnostic code in CDARS was validated with a positive predictive value (PPV) of 81.5% (95% confidence interval=76.1%-86.9%)(17).

The inclusion criteria included adult patients (age at or above 18 years old) with principal diagnosis of COPD identified by International Classification of Diseases, 9th Revision (ICD-9) code of 496 in the hospitalization from year 2022 to 2024. Patients with co-existing ICD-9 code of 493, which suggested co-existing diagnosis of asthma were excluded. The demographics (age and gender) and clinical characteristics (Charlson co-morbidity index (CCI), baseline blood eosinophil count (BEC), and medications for COPD) were retrieved from CDARS. The study was approved by the Institutional Review Board of the University of Hong Kong and Hospital Authority Hong Kong West Cluster (IRB/REC Reference No. UW 23-356). Patient informed consent was waived as it was a retrospective study without active patient recruitment, and all retrieved clinical data were de-identified.

AECOPD was defined as an event characterized by dyspnea and/or cough and sputum that worsens over ≤14 days, which may be accompanied by tachypnea and/or tachycardia, and is often associated with increased local and systemic inflammation caused by airway infection, pollution, or other insult to the airways, which is characterized by having an acute change in at least one of the following symptoms: cough increases in frequency and severity, sputum production increases in volume and/or changes character, dyspnea increases (18, 19). Mild exacerbation is defined as those treated with short acting bronchodilators (SABD) only. Moderate exacerbation is defined as those treated with SABDs and oral corticosteroids with or without antibiotics. Severe exacerbation is defined as patient that requires hospitalization or visits the emergency room (18).

The direct healthcare costs were estimated by the total healthcare costs attributed to emergency department visits, acute and convalescent medical ward admissions, as well as high dependency/intensive care unit admissions. The relevant daily costs were retrieved from Hospital Authority Statistical Report (20).

The primary outcome was the total head count and admission numbers related to severe AECOPD; as well as the direct healthcare costs associated with severe AECOPD. The secondary outcomes included factors associated with an increase in direct healthcare costs resulting from severe AECOPD.

### Statistical analysis

Categorical variables were expressed as frequency (percentage) and compared with Chi-squared tests or Fisher’s exact tests where appropriate. Continuous variables were expressed as mean ± standard deviation (SD) or median (25th to 75th centile) and compared with Student’s test or Mann Whitney test where appropriate. The number of severe AECOPD admissions over the 3 years were compared by one-way ANOVA. The relationship between disease factors and direct healthcare costs was assessed using Pearson’s correlation coefficients. Two-sided tests were performed, with statistical significance determined at the level of p < 0.05. All statistical analyses were performed using the 29th version of the SPSS statistical package and R version 4.4.1.

## Results

From 2022 to 2024, there were 11,465 COPD patients admitted to acute medical hospitals for severe AECOPD, contributing to 25,053 hospital admissions and 150,717 bed days. The total direct healthcare costs were HKD$ 1.259 billion for these severe AECOPD. 3,624 patients died within the study period. There were 10,421 (90.9%) male patients with mean age of 77.4 ± 10.2 years. The baseline demographics of the patients were illustrated in Table 1.

**Table 1.** Baseline clinical characteristics of patients with severe chronic obstructive pulmonary disease exacerbation.

|  | **Whole cohort**<br>**(n =11465)** |
| --- | --- |
| Age (years) | 77.4 ± 10.2 |
| Gender |  |
| Male | 10421 (90.9%) |
| Female | 1044 (9.1%) |
| CCI | 4.5 ± 1.8 |
| Severe AECOPD in prior 12 months | 430 (3.8%) |
| Co-morbidities |  |
| Hypertension | 4842 (42.2%) |
| Diabetes mellitus | 2004 (17.5%) |
| Hyperlipidaemia | 2394 (20.9%) |
| Stroke/TIA | 1019 (8.9%) |
| Heart failure | 1938 (16.9%) |
| Ischemic heart disease | 2021 (17.6%) |
| Bronchiectasis | 462 (4.0%) |
| Medication |  |
| ICS | 7891 (68.8%) |
| LABA | 9593 (83.7%) |
| LAMA | 9890 (86.3%) |
| Roflumilast | 1633 (14.2%) |
| Theophylline | 1920 (16.7%) |
| Baseline laboratory parameters |  |
| Eosinophil count (x cells/μL) | $452 \pm 589$ |
| Serum Cr (μmol/L) | $93.8 \pm 60.1$ |
| Serum albumin level (g/L) | $33.6 \pm 10.2$ |

TIA, transient ischaemic attack; ICS, inhaled corticosteroids; LABA: long-acting beta-agonist;
LAMA: long-acting anti-muscarinic; Cr, creatinine
Data expressed as mean ± S.D.

### Trends from 2022 to 2024

In the year 2022, there were 4,025 patients admitted to acute medical hospitals for severe AECOPD, with a total of 6,178 admissions. In the year 2023, there were 6,095 patients admitted to acute medical hospitals for severe AECOPD, with a total of 8,754 admissions. In the year 2024, there were 6,374 patients admitted to acute medical hospitals for severe AECOPD with a total of 10,121 admissions. The p-value between-group differences for the number of severe AECOPD admissions was 0.003, suggesting a significant increase in the severe AECOPD admission numbers over 3 years (Figure 1).

**Figure 1.**
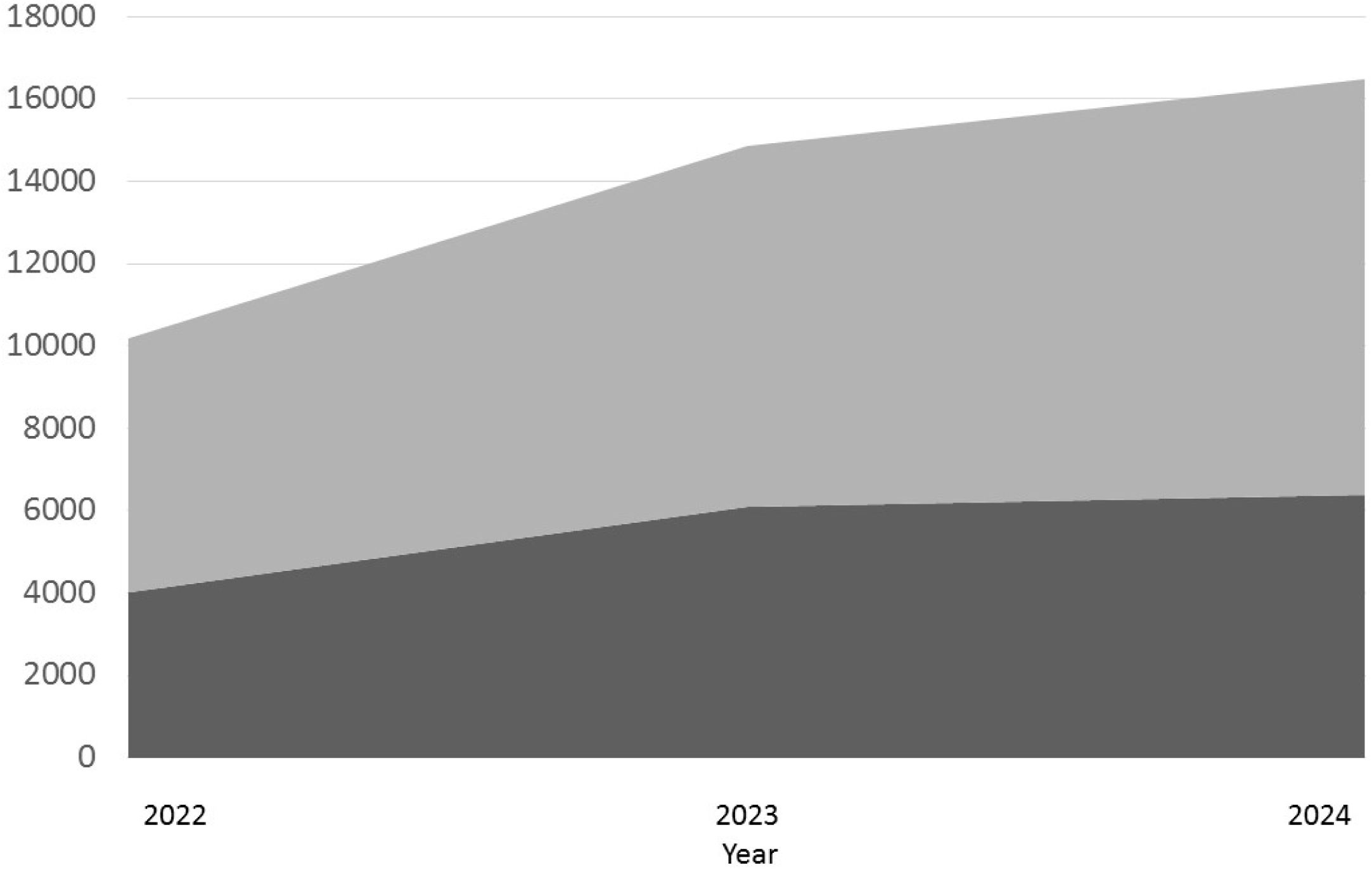
Number of headcount admitted for severe AECOPD and number of severe AECOPD hospital admission from year 2022 to 2024.

### Phenotype and burden of AECOPD

Among these patients, 5,128 (44.7%) of them had eosinophilic phenotype with baseline BEC ≥ 300 cells/µL, while 6,337 (55.3%) had non-eosinophilic phenotype with baseline BEC < 300 cells/µL.

The mean annual total healthcare costs related to severe AECOPD was HKD$ 79,230 ± 131,450 for patients with non-eosinophilic phenotype and HKD$ 80,136 ± 117,608 for patients with eosinophilic phenotype, p = 0.70 (Figure 2).

**Figure 2.**
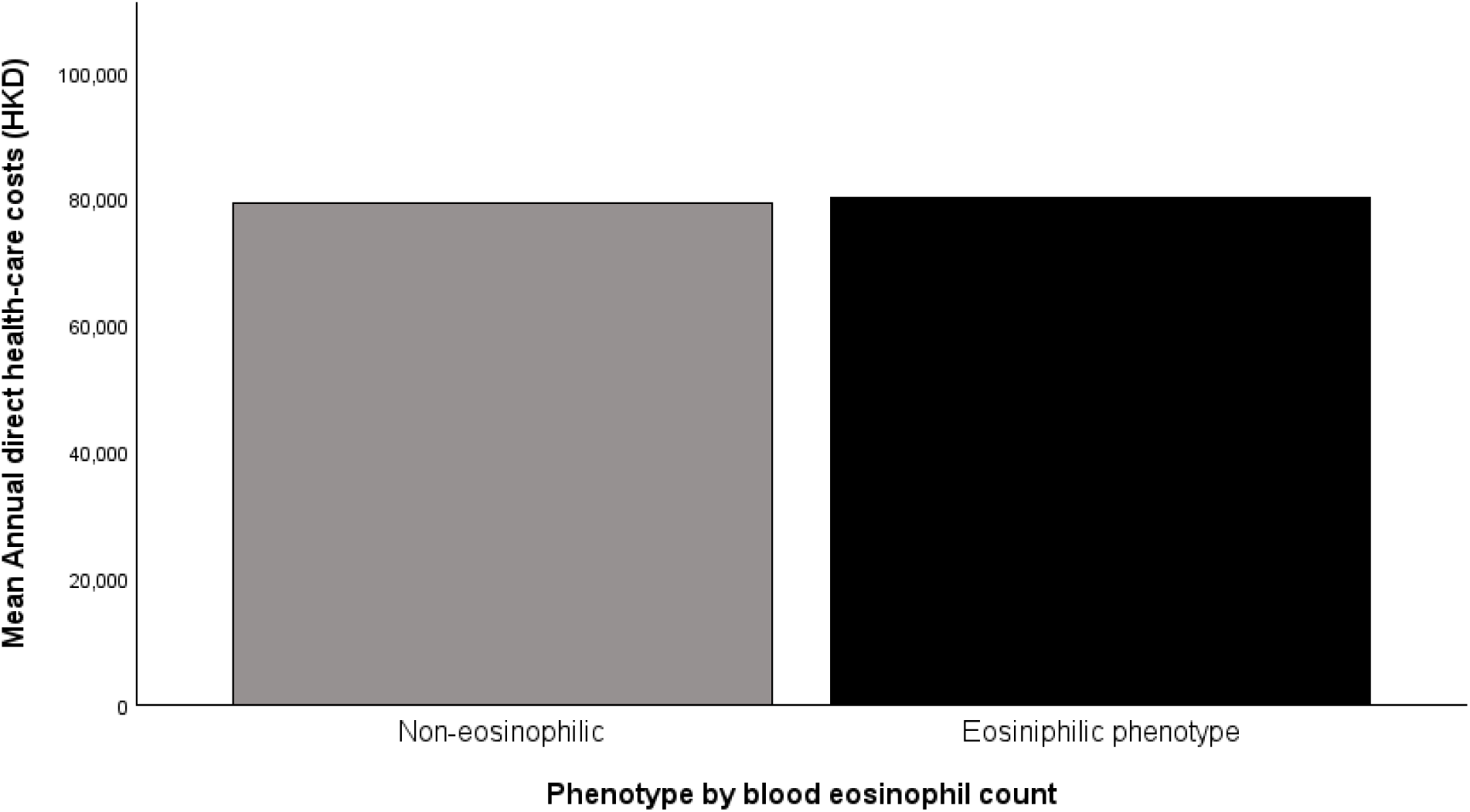
Annual total direct healthcare costs by phenotype.

Patients with co-existing bronchiectasis (n = 462) had a significantly higher annual number of severe AECOPD and annual direct healthcare costs than those without co-existing bronchiectasis (n = 11,003) (Annual number of severe AECOPD: 2.16 ± 2.84 vs 1.49 ± 1.61, p < 0.001; HKD$ 113,412 ± 145,593 vs HKD$ 78,218 ± 124,332, p < 0.001)

### Exacerbation number and burden of AECOPD

Among the included patients, the mean annual number of severe AECOPD was 1.52 ± 1.68 times per year. There were 1,653 patients with ≥ 2 severe AECOPD per year and 9,812 with < 2 severe AECOPD per year. The mean annual total healthcare costs related to severe AECOPD was HKD$ 55,309 ± 84,334 for patients with < 2 severe AECOPD per year and HKD$ 224,037 ± 206,351 for patients with≥ 2 severe AECOPD per year, p < 0.001. The median annual total healthcare costs related to severe AECOPD was HKD$ 30,148 (15,074–62,273) for patients with < 2 severe AECOPD per year and HKD$ 162,478 (90,444–288,043) for patients with ≥ 2 severe AECOPD per year, p < 0.001 (Figure 3). The annual number of severe AECOPD significantly correlates with total direct healthcare costs and annual direct healthcare costs related to severe AECOPD with r = 0.536, p < 0.001 and r = 0.612, p < 0.001, respectively.

**Figure 3.**
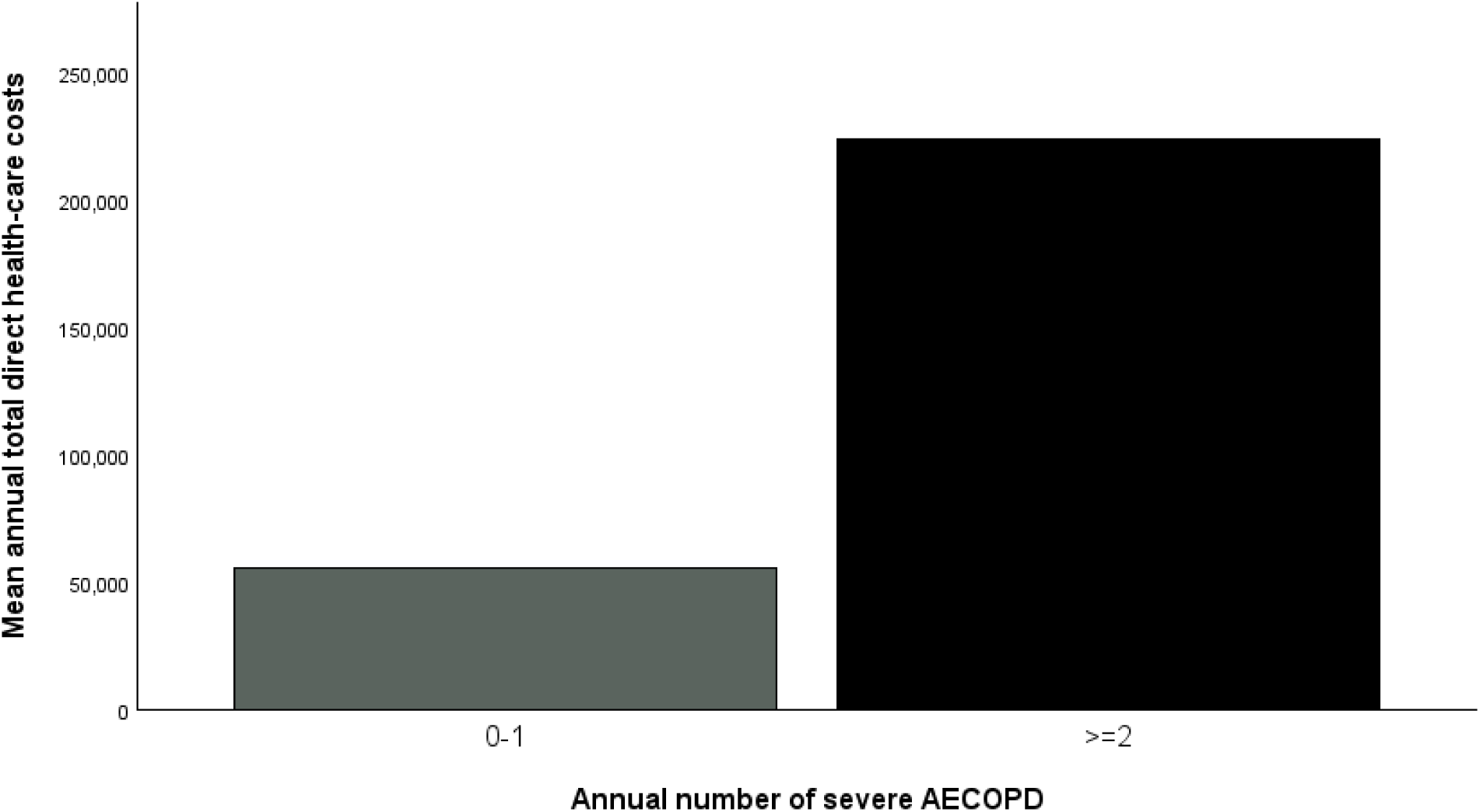
Annual total direct healthcare costs by annual severe AECOPD number.

The annual number of severe AECOPD in the 12-month period prior was significantly associated with subsequent annual number of severe AECOPD and the associated direct healthcare costs with r = 0.553, p < 0.001 and r = 0.355, p < 0.001 respectively.

## Discussion

Our study illustrated the healthcare and economic burden of severe AECOPD in Hong Kong in the post-COVID era. Severe AECOPD remains a major burden leading to significant healthcare utilization due to the consumption of medical beds, as well as the associated direct healthcare costs. We also observed the increasing number of severe AECOPD from 2022 to 2024, suggesting the post-pandemic effect, which was also observed in other diseases(21, 22, 23). The post-pandemic changes reported also involve the changes in respiratory pathogen epidemiology, which could influence the severe AECOPD number, as bacterial and viral pathogens were estimated to be the culprit in causing AECOPD in up to 80% of the cases(24). The rising total headcount and admission numbers with the resulted direct healthcare costs lead to a major impact on healthcare burden.

In our study, we also demonstrated that severe AECOPD contributed significant direct healthcare costs, especially among patients with more frequent severe AECOPD. Co-existing bronchiectasis is also another factor associated with increased direct healthcare costs. Regarding the phenotype as stratified by blood eosinophil count, both eosinophilic and non-eosinophil groups carry equally high direct healthcare costs. These findings once again echoed the importance of severe AECOPD as the important contributor to healthcare and economic burden (25). The finding also concurred that the presence of co-existing diseases and comorbidities is the factor associated with the healthcare and economic burden (26). Among all the co-existing diseases and comorbidities, bronchiectasis is common and has also been reported to have dynamic interaction with COPD, resulting in an increase in healthcare and economic burden. Bronchiectasis was reported to be present in 5 to 10% of the patients with AECOPD.(27, 28) Kim et al, reported that patients with COPD and bronchiectasis to have higher rate of acute exacerbation requiring antibiotics, moderate-to-severe exacerbation, as well as higher treatment cost and duration than those with COPD without bronchiectasis (29). COPD patients with bronchiectasis were also reported to require longer hospitalization during exacerbation (27). However, such phenomenon on the negative impact of bronchiectasis on AECOPD was also challenged by other studies (28).

The important healthcare and economic burden of severe AECOPD call for strategies in preventing AECOPD. In the 2025 version of GOLD report(30), two new drugs have been included as add-on therapy for patients with Group E COPD. These include interleukin (IL)-4 receptor (alpha subunit) monoclonal antibody dupilumab and phosphodiesterase (PDE) 3 and 4 Inhibitor ensifentrine (31, 32, 33, 34). GOLD also recommended vaccinations against influenza, severe acute respiratory syndrome coronavirus 2 (SARS-CoV-2), Streptococcus pneumoniae, respiratory syncytial virus (RSV), pertussis and varicella zoster (VZV)(30), while bacteria and viruses are well reported to be trigger for AECOPD. Timely and appropriately initiate strategies to prevent AECOPD should be initiated given the negative impact from AECOPD.

Another important finding is that 3,624 out of 11,465 (28.5%) patients died within the study period. This is an alarming observation which was also reported in prior studies. Slenter et al. reported that the one-year mortality rate after a AECOPD was 28%(35). The one-year mortality rate was even higher among those who required invasive or non-invasive ventilation during their hospitalization, at 45.7% and 41.8% respectively (36). Our findings, together with what was reported in the literature, highlighted the risks associated with AECOPD beyond the readmission and healthcare burden, but also an alarming mortality rate.

Taken together, severe AECOPD remains an important healthcare issue in Hong Kong with a trend towards a rising burden after COVID-19 pandemic. Severe AECOPD not only resulted in bed utilization but also huge economic costs from direct healthcare costs. Last but not least, the mortality after severe AECOPD is high as in literature and clinicians should not underestimate the potential harm of AECOPD.

Our study has several limitations. Firstly, we have not analyzed the granular details of etiology of the severe AECOPD episodes. But as the aim of this study is to analyze the whole picture of severe AECOPD burden in Hong Kong, these details may not have major implications here. Secondly, the majority of the patients included in this study were Chinese, which may affect the generalizability. But we also reported similar findings as mentioned in the sessions before which suggested this ethnic difference may not be significant. Thirdly, lung function parameters and symptom burden were not assessed here. But this may not affect the overall results as the aim of this study is on severe AECOPD and its burden. Lastly, drug costs and income loss were not included in the calculation of economic burden. But all the patients in this study were having GOLD Group E COPD and the various inhalers in Hong Kong have similar pricing. Most of the patients included were beyond retirement age and the income loss is considered to be negligible.

### Conclusions

Severe AECOPD carries significant burden towards healthcare system. Patients with bronchiectasis and more frequent severe AECOPD were factors associated with the higher number of severe AECOPD episodes and the associated direct healthcare costs.

## Author Contributions

Kwok WCH, Ali C: Conception and design of study, analysis of data, writing of manuscript; Wong CK, Leung ISH Ho JCM: Revision of manuscript, data curation, supervision, Ma TF, Zhou L: data analysis and processing.

## Funding

This research was supported by Sanofi Hong Kong.

## Data Availability Statement

All available data are presented in the manuscript and no additional data will be provided.

## Conflicts of interest

The authors declare no conflict of interest.

## Notes

### Competing Interest Statement

The authors have declared no competing interest.

### Author Declarations

The study was approved by the Institutional Review Board of the University of Hong Kong and Hospital Authority Hong Kong West Cluster (IRB/REC Reference No. UW 23-356).

